# Magnitude and breadth of neutralizing antibody responses elicited by SARS-CoV-2 infection or vaccination

**DOI:** 10.1101/2021.12.30.21268540

**Authors:** Benjamin L. Sievers, Saborni Chakraborty, Yong Xue, Terri Gelbart, Joseph C. Gonzalez, Arianna G. Cassidy, Yarden Golan, Mary Prahl, Stephanie L. Gaw, Prabhu S. Arunachalam, Catherine A. Blish, Scott D. Boyd, Mark M. Davis, Prasanna Jagannathan, Kari C. Nadeau, Bali Pulendran, Upinder Singh, Richard H. Scheuermann, Matthew Frieman, Sanjay Vashee, Taia T. Wang, Gene S. Tan

**Author notes:** these authors contributed equally (Benjamin L. Sievers, Saborni Chakraborty, Yong Xue, Terri Gelbart). these authors contributed equally (Sanjay Vashee, Taia T. Wang, Gene S. Tan).

## Abstract

Multiple SARS-CoV-2 variants that possess mutations associated with increased transmission and antibody escape have arisen over the course of the current pandemic. While the current vaccines have largely been effective against past variants, the number of mutations found on the Omicron (B.1.529) spike appear to diminish the efficacy of pre-existing immunity. Using pseudoparticles expressing the spike of several SARS-CoV-2 variants, we evaluated the magnitude and breadth of the neutralizing antibody response over time in naturally infected and in mRNA-vaccinated individuals. We observed that while boosting increases the magnitude of the antibody response to wildtype (D614), Beta, Delta and Omicron variants, the Omicron variant was the most resistant to neutralization. We further observed that vaccinated healthy adults had robust and broad antibody responses while responses were relatively reduced in vaccinated pregnant women, underscoring the importance of learning how to maximize mRNA vaccine responses in pregnant populations. Findings from this study show substantial heterogeneity in the magnitude and breadth of responses after infection and mRNA vaccination and may support the addition of more conserved viral antigens to existing SARS-CoV-2 vaccines.

**One Sentence Summary:** Diminished efficacy of pre-existing immunity to highly mutated SARS-CoV-2 variants.

## INTRODUCTION

First identified in Botswana in November 2021, the SARS-CoV-2 Omicron variant (B.1.1.529) is rapidly becoming the dominant circulating variant of concern (VOC)(*1, 2*). The Omicron virus harbors a striking 59 amino acid substitutions throughout its genome relative to the ancestral Wuhan-hu-1 SARS-CoV-2 virus, referred to as D614 here. Thirty-seven of these mutations are within the spike protein, the target of neutralizing antibody responses against this virus. As neutralizing antibodies are the major correlate of protection against coronavirus disease 2019 (COVID-19)(*3, 4*), this degree of mutational change raises questions about the effectiveness of neutralizing antibodies that were elicited by infection with SARS-CoV-2 D614 infection or by current mRNA vaccines which encode the wildtype (WT) spike. To define the extent of escape by Omicron from neutralizing antibodies in the population, we evaluated the magnitude and breadth of the response against the D614 WT virus along with three VOCs, Beta (B.1.351), Delta (B.1.617.2) and Omicron (B.1.1.529). Understanding these neutralizing antibody responses will enable us to assess the state of pre-existing immunity elicited by the WT virus and can inform the design of the next generation of COVID-19 vaccines.

The spike glycoprotein of SARS-CoV-2 has two major antigenic domains; mutations in these regions can contribute to antigenic escape and reduced immunity against infection(*5*). The receptor binding domain (RBD) interacts directly with the receptor for SARS-CoV-2, angiotensin-converting enzyme 2 (ACE2), and amino acid changes in RBD can impact the affinity of spike for ACE2 and thus transmissibility and virulence of viral variants. The Beta variant has notable mutations (L18F, D80A, D215G, Δ242-244 and R246L) in the amino terminal domain (NTD) and RBD (K417N, E484K and N501Y) (*6*) of the spike protein that is associated with antibody escape as previously reported (*7-9*). The emergence of the Delta with changes in the RBD resulted in higher transmissibility and ultimately has become the predominant circulating strain of SARS-CoV-2 currently. Unlike Beta, Delta has only two mutations in the RBD (L452R, E484Q) relative to the WT virus that are associated with antibody escape(*4*). Based on these different mutational profiles of the Beta and Delta spike proteins, we chose to include these VOCs in the present study along with Omicron. Omicron harbors a relative abundance of mutations with 37 non-synonymous changes in the spike alone, 11 in the NTD and 15 in the RBD. Based on the structural features of the Omicron spike and recent findings by other groups (*10-13*), we anticipated that Omicron would be at least as resistant to current neutralizing antibodies in the population as the Beta variant and likely far more resistant compared to the WT and Delta viruses.

To study the relative susceptibility of the spikes of SARS-CoV-2 VOC to neutralizing antibodies in the population, we studied activity in sera or plasma from three cohorts of naturally infected or mRNA-vaccinated individuals against D614, Beta, Delta and Omicron VOCs. In a natural infection cohort, we tested plasma collected during the peak phase of mild COVID-19 (day 28 post study enrollment) and two time points during the convalescent period – at days 210 and 300 post study enrollment. To understand the breadth of neutralizing antibodies elicited by mRNA vaccination, we studied a cohort of pregnant individuals who received two doses of the Pfizer or Moderna vaccines during pregnancy; pregnancy is a risk factor for poor outcomes in COVID-19, thus this cohort provides important insights into immunity in this vulnerable population. A second cohort of healthcare workers received three doses of the Pfizer vaccine. These approved SARS-CoV-2 mRNA vaccines code for the original D614 SARS-CoV-2 spike protein. While mRNA vaccines have been extremely successful at inducing potent neutralizing antibody responses, their effectiveness will depend in large part to the degree of antigenic drift in circulating SARS-CoV-2 variants. Using these three distinct cohorts, we evaluated the magnitude and breath of neutralizing antibody titers over time after natural infection and mRNA vaccination.

## RESULTS

### Magnitude and breadth of the antibody response following natural infection

We first evaluated a total of 54 plasma samples from three study time-points for neutralizing antibody responses in a group of individuals from a longitudinal cohort of mild SARS-CoV-2 patients enrolled in an outpatient study at Stanford Hospital Center (*14*). This study was a trial evaluating the efficacy of interferon lambda in mild COVID-19 but only subjects from the placebo arm have been studied here. Subjects in this study were infected with SARS-CoV-2 during the first half of 2020, a time when the D614 virus was the dominant circulating SARS-CoV-2 strain. Three time points were chosen for this analysis: day 28 post enrollment, the previously characterized peak antibody response, and days 210 and 300 post enrollment (**Figure 1A**) (*15*). Neutralizing antibody titers were highest on day 28 for all VOCs and waned over time in the convalescent period on days 210 and 300 (**Figure 1B**). Notably, both Beta and Omicron VOC were much more resistant to neutralization even on day 28 compared to WT and Delta, perhaps reflecting the key mutations found in both the Beta and Omicron spike that have been previously described to contribute to antibody escape (*8*). While neutralizing activity against D614 and Delta was measurable in many subjects at day 300, activity against the Beta and Omicron variants was largely absent by this later timepoint (**Figures 1B, C, D**).

**Figure 1.**
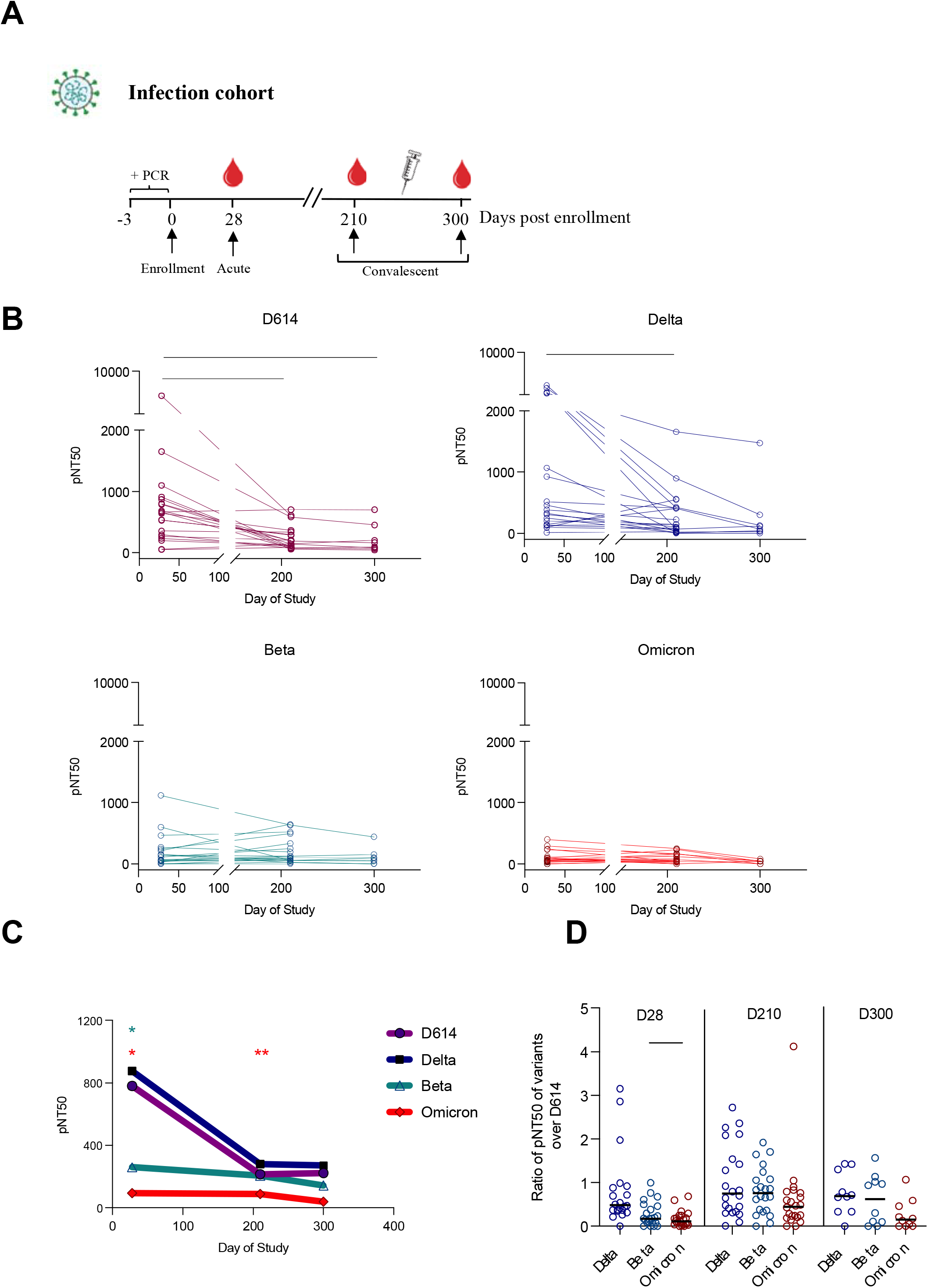
Plasma neutralizing titers against variants of concern in an outpatient COVID-19 infection cohort. (A) Study subjects were enrolled (day 0) within three days of a positive SARS-CoV-2 PCR test. Longitudinal plasma samples at an early study time-point (day 28, n=23), and two convalescent time points (day 210, n=23 and day 300, n=8) were assessed for neutralizing antibody against 4 SARS-CoV-2 pseudovirus strains: D614, Delta (B.1.617.2), Beta (B.1.351) and Omicron (B.1.1.529). (B) The kinetics of half-maximal SARS-CoV-2 pseudovirus neutralizing titers (pNT50) over time against D614, Delta (B.1.617.2), Beta (B.1.351) and Omicron (B.1.1.529). (C) Mean pNT50 values across study time-points of all 4 pseudovirus variants. Values significantly different from D614 at each time point are shown. (D) Ratio of pNT50 values of the variants of the concern Delta, Beta and Omicron over D614 pNT50 at each study time point. p values in (B-D) were calculated using mixed effects analysis with Geisser-Greenhouse correction and Tukey’s multiple comparisons test.

### Magnitude and breadth of the antibody response following vaccination

To understand the neutralizing antibody responses elicited by SARS-CoV-2 mRNA vaccines, we studied two separate cohorts of vaccinated individuals. One cohort comprised pregnant subjects that were enrolled in a vaccine study in the University of California San Francisco Health system (n=9 at baseline before vaccination and n=33 following 2 doses of mRNA vaccine)(*16, 17*). These subjects received either the Pfizer (n=10) or Moderna (n=23) mRNA vaccines during pregnancy. We tested paired neutralizing antibody responses in a subset of subjects (n=9) from serum taken before vaccination (baseline) and in serum collected after the second immunization (post-dose 2 (PD2)) (**Figure 2A**). As expected, the PD2 neutralizing titers were substantial against D614 (the homologous spike) after two vaccine doses, with a mean pNT50 of ∼128 fold over baseline. Response at PD2 in all subjects were progressively lower than D614, with neutralizing titers against Delta (91x)>Beta (40x)>Omicron (10.2x) (**Figure 2B**). The magnitude of neutralizing titers against Delta, Beta and Omicron were all significantly reduced relative to D614 (**Figure 2C**). Substantial heterogeneity was present among vaccinees in the breadth of neutralizing antibody responses. This was evidenced in the wide range of ratios displayed by individuals in neutralizing titers against Delta, Beta and Omicron variants relative to D614 (**Figure 2D**).

**Figure 2:**
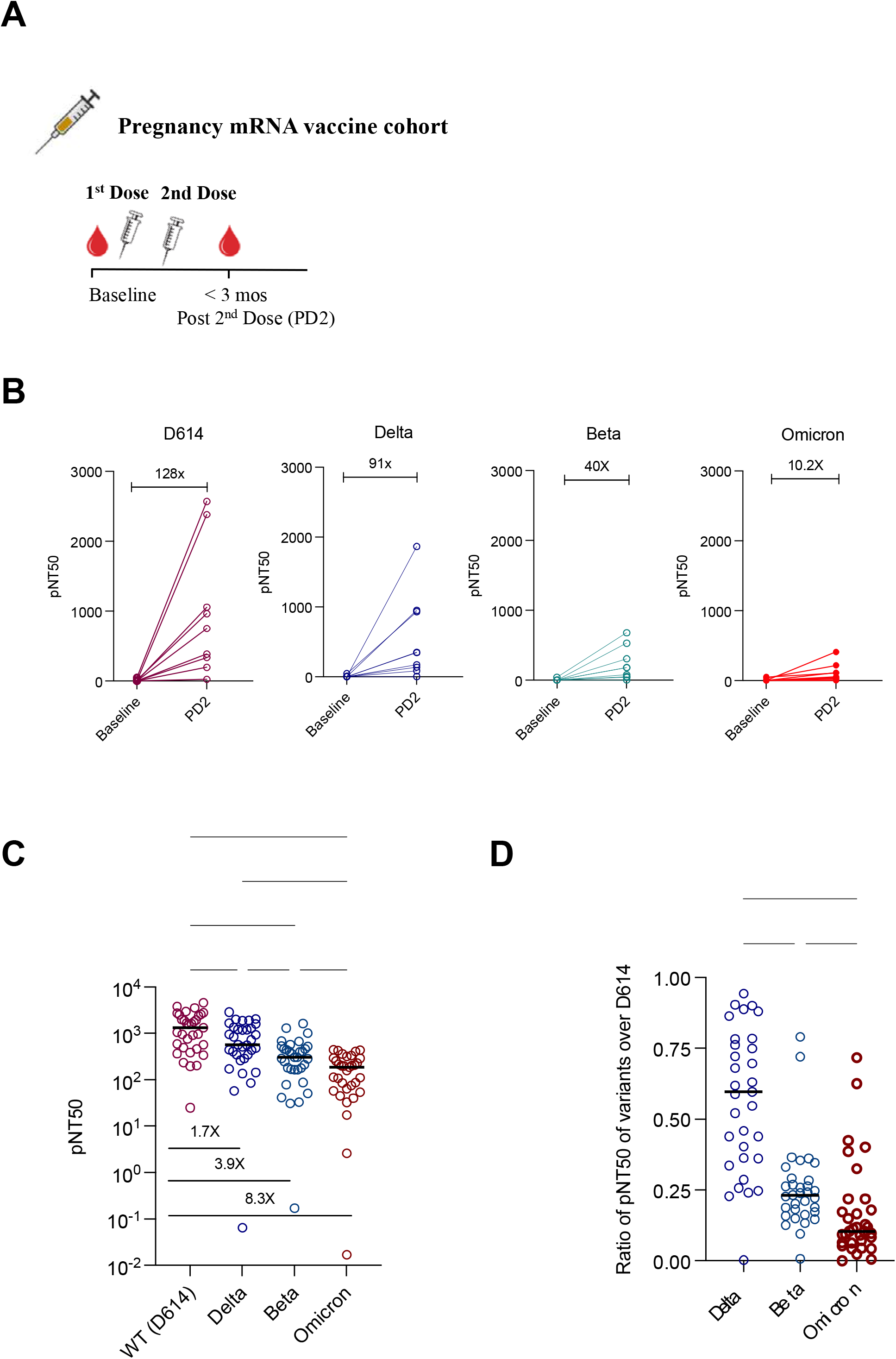
Plasma neutralizing titers against variants of concern in a mRNA vaccinated cohort of pregnant women. **(A)** Samples collected from a cohort of vaccinated pregnant women at baseline (n=9) and after 2 doses of mRNA vaccine (n=33) were assessed for neutralizing antibody against 4 SARS-CoV-2 pseudovirus strains: D614, Delta (B.1.617.2), Beta (B.1.351) and Omicron (B.1.1.529). (B) Half-maximal SARS-CoV-2 pseudovirus neutralizing titers (pNT50) against D614G, Delta (B.1.617.2), Beta (B.1.351) and Omicron (B.1.1.529) of paired samples at baseline and 2 doses of mRNA vaccine. Fold change of mean titers between the 2 time-points are shown. (C) pNT50 values of samples collected after 2 doses of mRNA against the 4 SARS-CoV-2 pseudoviral variants. Fold reduction of mean pNT50 compared to D614G are shown for each variant. (D) Ratio of pNT50 values of the variants of the concern Delta, Beta and Omicron over D614 pNT50. p values in (C-D) were calculated using mixed effects analysis with Geisser-Greenhouse correction and Tukey’s multiple comparisons test.

Finally, we evaluated the neutralizing antibody responses from a cohort of healthcare workers previously vaccinated with the Pfizer/BNT162b2 mRNA vaccine at the Stanford Hospital Center (n=137 samples from 4 study timepoints)(*18, 19*). Four time points were chosen for this analysis. An early timepoint following the second dose (seven days post dose 2 or study day 28), followed by a late timepoint at study day 210 prior to dose 3 enabled us to study the durability of neutralizing responses after 2 doses of vaccine. In addition, we defined neutralizing responses on day 7 or between day 21-28 post dose 3 (**Figure 3A**). Neutralizing antibody titers were generally highest at the timepoints 7 days after dose 2 and dose 3 against the homologous D614 and all variants. As expected, titers had waned substantially against all variants by day 210 after dose 2. The third vaccine dose increased the titers against all variants substantially, to a level that matched, and in most cases surpassed, titers observed at 7 days post dose 2 (**Figure 3B**). By the day 21-28 post dose 3 timepoint, some individuals already had reduced neutralizing antibody levels while others maintained durable levels approximating those observed on 7 days post dose 3. The antibody titers against Omicron were measurable in all subjects after dose 3 but were significantly lower than other variants (**Figures 3C and D**). As in the pregnancy cohort, substantial heterogeneity was observed in the breadth of the neutralizing antibody responses, evidenced by the ratio of titers against variants to D614 (**Figures 3C and D**).

**Figure 3:**
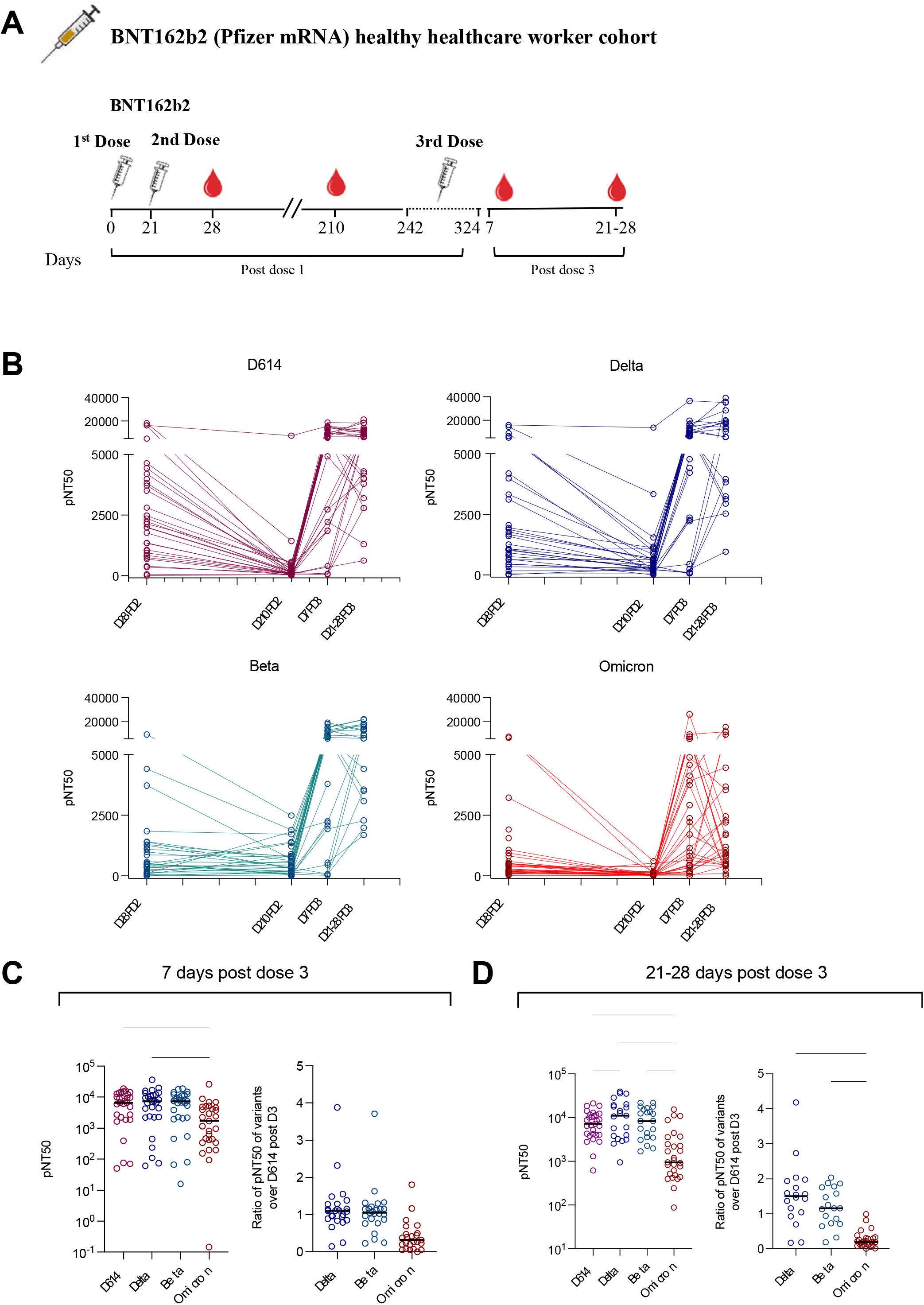
Plasma neutralizing titers against variants of concern in vaccinated cohort of healthcare workers who received 3 doses of mRNA vaccine. **(A)** Samples were collected from a cohort of healthcare workers who received 2 doses of Pfizer vaccine 21 days apart and received a 3^rd^ dose 242-324 days after the first dose. Antibody titers were assessed at 28 and 210 days after the first dose, and 7 days and 21-28 days after the third. **(B)** The kinetics of half-maximal SARS-CoV-2 pseudovirus neutralizing titers (pNT50) over various study time points against D614, Delta (B.1.617.2), Beta (B.1.351) and Omicron (B.1.1.529). pNT50 values (left panel) and ratio of pNT50 of the variants of the concern Delta, Beta and Omicron over D614 pNT50 (right panel) of samples collected **(C)** 7 days after 3^rd^ dose of vaccine and **(D)** 21-28 days after 3^rd^ dose of vaccine. p values in (C-D) were calculated using ordinary one-way ANOVA with Tukey’s multiple comparisons test.

## DISCUSSION

Using three longitudinal cohorts, we measured the magnitude and breath of the antibody response against WT, Delta, Beta and Omicron SARS-CoV-2 viruses following natural infection or mRNA vaccination. Infection during the early pandemic when D614 was the dominant circulating SARS-CoV-2 virus elicited a relatively low neutralizing antibody response against Beta and Omicron variants while immunity against Delta was not significantly reduced over D614. There was significant heterogeneity in this cohort with respect to both the magnitude and breadth of neutralizing responses against all variants tested. In two vaccine cohorts, we observed robust responses elicited by mRNA SARS-CoV-2 vaccines. In all three cohorts, responses against Beta and Omicron were reduced relative to the homologous D614 and Delta, as has been previously described (*5-7*). In a cohort of healthcare workers where we were able to evaluate responses after 3 vaccine doses, there was clearly a benefit to both magnitude and breadth of the response conferred by the 3^rd^ dose. Most importantly, titers against Omicron were robust after dose 3 relative to dose 2. There was substantial heterogeneity in responses, particularly with respect to breadth, after 2 and 3 doses of mRNA vaccines. Determinants of broadly neutralizing antibody responses after homologous vaccine boosts are not well understood at this time and are an important topic to address in future studies.

Because COVID-19 is a risk factor for adverse outcomes in pregnancy, it is critical to understand the response to mRNA vaccines in this population. Here, we show a relatively consistent response in the magnitude of response after two vaccine doses in this population but note that, while timepoints were not exactly matched with those in the healthy vaccine cohort, the titers were generally lower in pregnancy. Understanding optimal timing during pregnancy for booster doses will be important for protecting this population(*20*).

Most approved vaccines rely solely on eliciting immune responses against a single immunogen, the spike. While clinical trials have demonstrated excellent effectiveness of mRNA vaccines up to now, it is evident that they will likely have somewhat reduced effectiveness against variants that are substantially antigenically drifted such as Omicron. As SARS-CoV-2 evolves, the efficacy of vaccines that relies on eliciting responses against a highly mutable antigen will always come into question. While it is noted that T-cell epitopes are generally not highly impacted by changes seen in the variants (*21*) as B cell epitopes we expect that supplementing spike-based vaccines with other more conserved viral antigens would elicit more potent and broad immunity against SARS-CoV-2 infections and COVID-19 (*22*). The role of non-neutralizing antibodies and T cells in immunity against VOCs warrants further study to better define all immune mechanisms, in addition to neutralizing antibodies, that can control viral replication (*23-26*)

## MATERIALS AND METHODS

### Clinical cohorts and samples

Characterization of these samples at Stanford was performed under a protocol approved by the Institutional Review Board of Stanford University (protocol #55718).

#### Stanford Lambda cohort

120 participants were enrolled in a phase 2 randomized controlled trial of Peginterferon Lambda-1a (Lambda, NCT04331899) Inclusion/exclusion criteria and the study protocol for the trial have been published(*12*). Briefly, adults aged 18-75 years with an FDA emergency use authorized reverse transcription-polymerase chain reaction (RT-PCR) positive for SARS-CoV-2 within 72 hours prior to enrollment were eligible for study participation. Exclusion criteria included hospitalization, respiratory rate >20 breaths per minute, room air oxygen saturation <94%, pregnancy or breastfeeding, decompensated liver disease, recent use of investigational and/or immunomodulatory agents for treatment of COVID-19, and prespecified lab abnormalities. All participants gave written informed consent, and all study procedures were approved by the Institutional Review Board of Stanford University (IRB-55619). Participants were randomized to receive a single subcutaneous injection of Lambda or saline placebo. Peripheral blood was collected at enrollment, day 5, and day 28 post enrollment. A subset of participants (n=80) returned for long-term follow-up visits 4-, 7-, and 10-months post enrollment, with peripheral blood obtained. Longitudinal samples from the 56 SARS-CoV-2-infected outpatients who were in the placebo arm of the broader Lambda study were obtained and assessed here.

#### Stanford Adult vaccine cohort

Fifty-seven healthy volunteers were enrolled in the study approved by Stanford University Institutional Review Board (IRB 8629). The median age was 36 years old with a range from 19 - 79 years old. There are 28 males and 29 females in the study. There are 27 White participants, 23 Asian participants, 4 Black participants, 1 Native American participant, and 2 other participants.

#### UCSF Pregnancy vaccine cohort

Fifty-eight female pregnant volunteers that received either the Moderna or Pfizer vaccine were enrolled in a study approved by the University of California at San Francisco Institutional Review Board (20-32077). Plasma samples were collected up to 24 hours before first vaccine dose, up to 24 hours before second dose, and 4-8 weeks after second vaccine dose. The median age at enrollment was 35 years old (range 27-42). The median gestational age was 21.5 weeks (range 5 to 40) (*16, 17*)

A subset of available plasma samples (as indicated in the main text) from all the cohorts were used in this study.

### SARS-CoV-2 spike variant spike construction

The WT SARS-CoV-2 spike gene was previously amplified with KOD Xtreme Hot Start DNA polymerase (Millipore) using cDNA from SARS-CoV-2/human/USA/WA-CDC-WA1/2020 (GenBank MN985325.1) and cloned into pCC1BAC-his3 vector (*27*). To generate SARS-CoV-2 variants’ spike genes, fragments were amplified with Platinum SuperFi II DNA polymerase (Thermo Fisher) using WT spike plasmid as a template. Desired mutations (Table 1) were introduced by primers to each amplicon which has 30-35 bp homologous sequences at each end to the adjacent fragments. These amplicons were digested with DpnI (NEB) to remove template DNA and purified by Qiagen PCR purification kit. 50 fmol of each amplicon and 15 fmol of YCP/BAC vector were covalently joined using standard Gibson assembly reaction (NEB), transformed into *E*.*coli* DH10B competent cells (Thermo Fisher), and plated on LB medium with 12.5 mg/ml chloramphenicol. *E*.*coli* transformants were verified to contain correct mutations using PCR and Sanger sequencing (GeneWiz). Plasmids were isolated from *E. coli* by the Purelink HiPure Plasmid Midiprep Kit (Thermo Fisher). Primers used for spike gene construction and verification are listed in Table 2. Lastly, the spike genes lacking the cytoplasmic domain by deleting the last 18 amino acides were then cloned into the pCAGGS expression vector.

**Table 1.** Variant spike mutations.

| Lineage | Spike AA substitutions vs WA1 |
| --- | --- |
| B.1.351 | L18F, D80A, D215G, R246I, K417N, E484K, N501Y, D614G, A701V |
| B.1.617.2 | T19R, G142D, E156-, F157-, R158G, L452R, T478K, D614G, P681R, D950N |
| B.1.529 | A67V, H69-, V70-, T95I, G142D, V143-, Y144-, Y145-, N211-, L212I, ins214EPE, G339D, S371L, S373P, S375F, K417N, N440K, G446S, S477N, T478K, E484A, Q493R, G496S, Q498R, N501Y, Y505H, T547K, D614G, H655Y, N679K, P681H, N764K, D796Y, N856K, Q954H, N969K, L981F, |

**Table 2.**
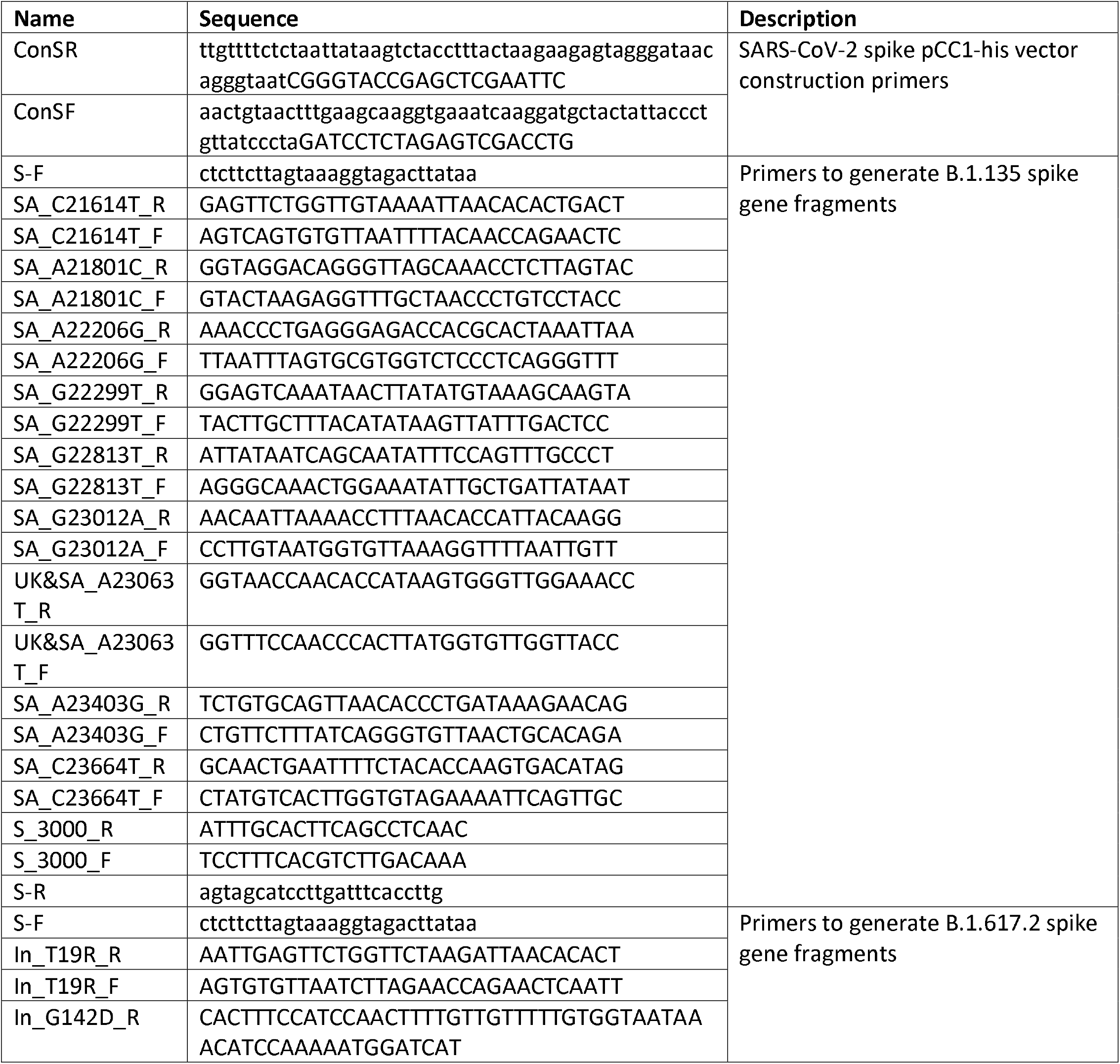
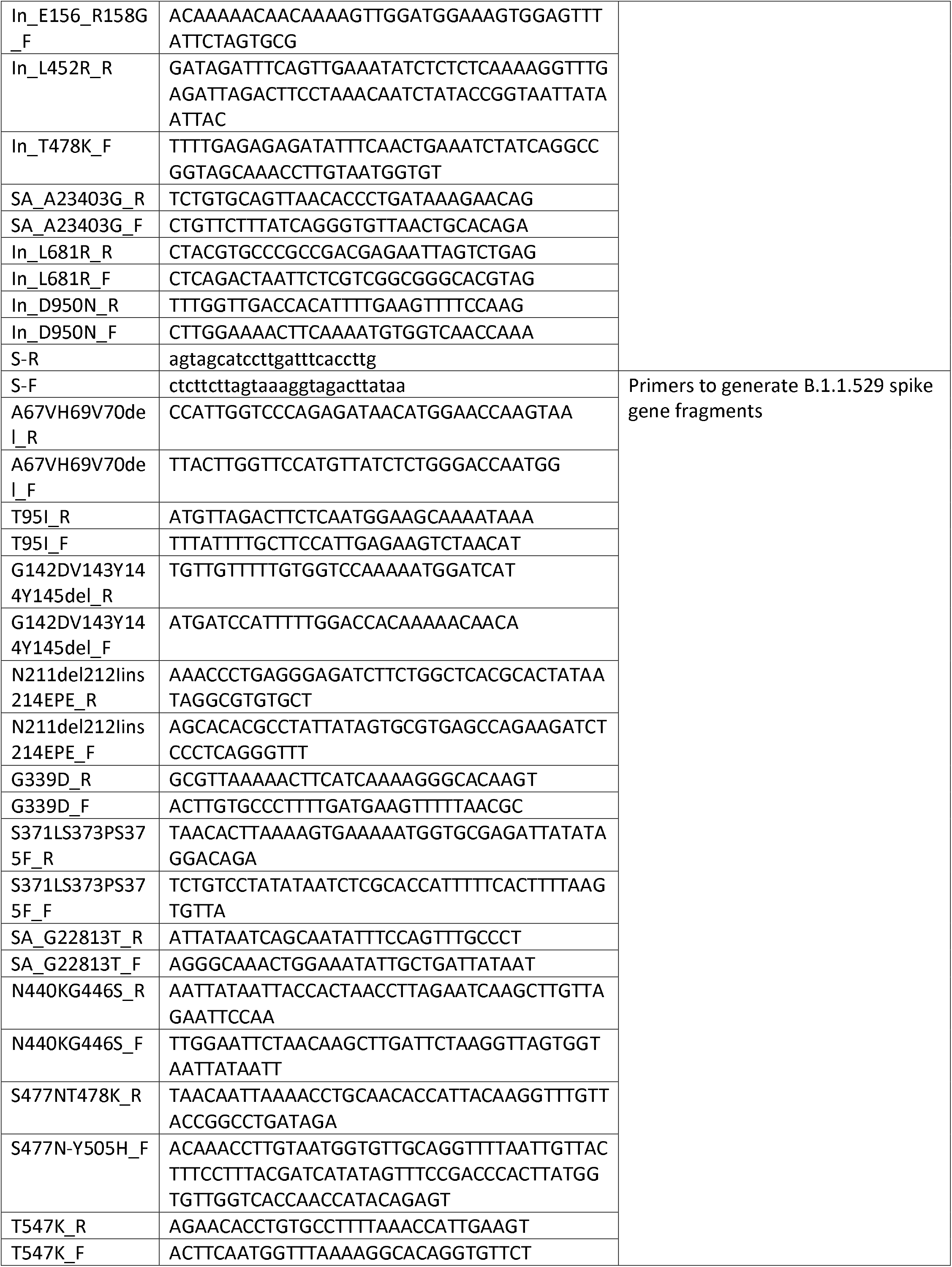
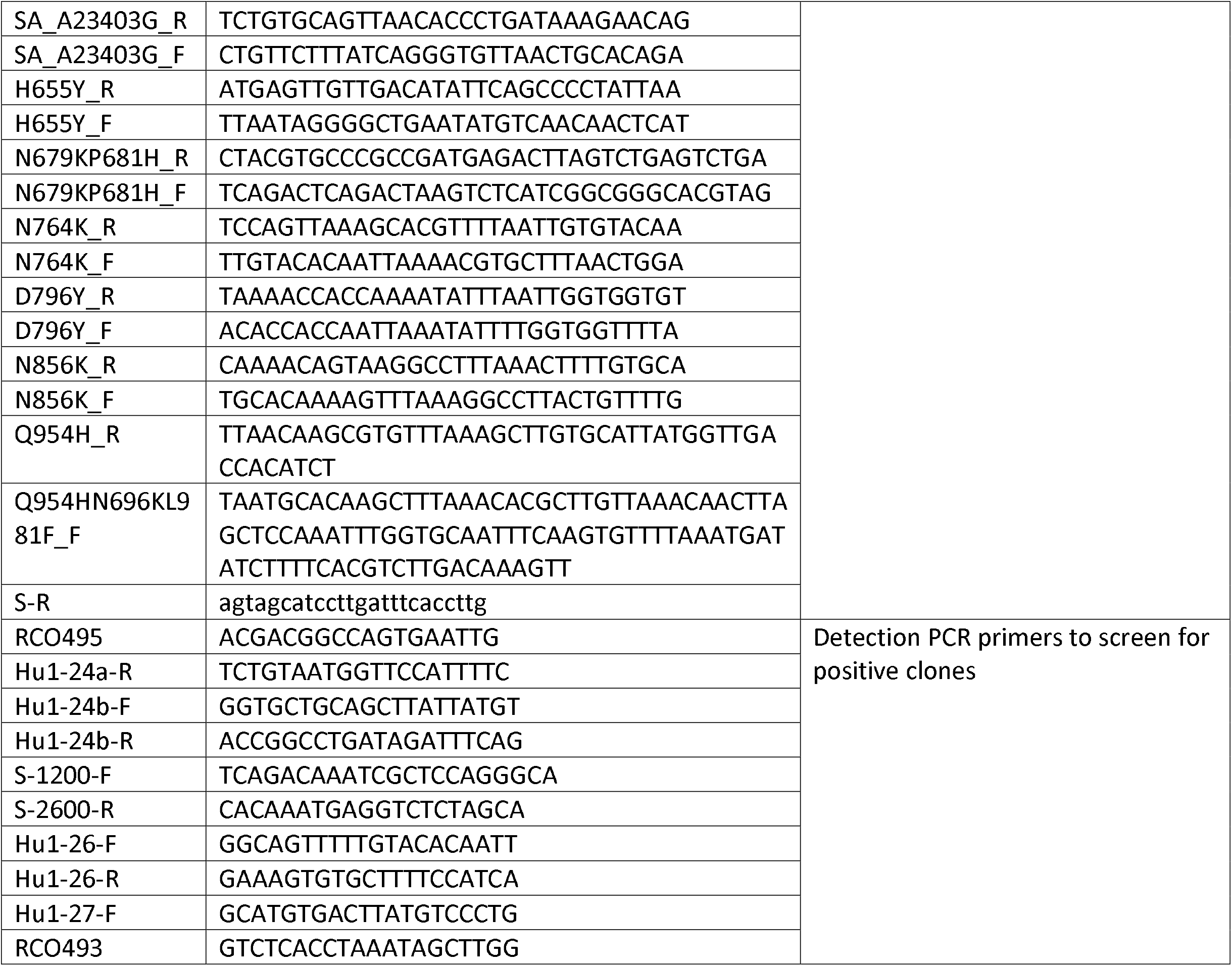
Primer sequences used for this study.

### Generation of SARS-CoV-2 pseudoparticles

To generate VSV pseudotyped with SARS-CoV-2 S, we first coated 6-well plates with 0.5 µg/mL poly-D-lysine (ThermoFisher, Cat. No. A3890401) for 1 to 2 hours at room temperature (RT). After poly-D-lysine treatment, plates were washed three times with sterile water and then seeded with 1.5e6 cells of HEK 293T per well. After 24 hours (h), cells were transfected with 1 µg of pCAGGS-SΔ18 per well using Lipofectamine 2000 transfection reagent (ThermoFisher, Cat. No., 11668019). 48 h after transfection, the cells were washed once with 1X phosphate buffered saline (PBS) and were infected with VSV-ΔG-GFP/nanoluciferase or VSV-ΔG-RFP/nanoluciferase (a generous gift from Matthias J. Schnell) at a multiplicity of infection of 2 to 3 in a 500 µL volume. Cells were infected for an hour with intermittent rocking every 15 minutes. After infection, the inoculum was carefully removed, and the cell monolayer was washed three times with 1X PBS to remove residual VSV-ΔG-GFP/nanoluciferase. Two mL of infection media (2% FBS, 1% glutamine, 1% sodium pyruvate in 1X DMEM) was added to each well. At 24 h post-infection, the supernatants from all the wells were combined and centrifuged (600 *g* for 30 min, 4°C), and stored at -80°C until use.

### Neutralization assays

Vero E6-TMPRSS2-T2A-ACE2 (obtained from BEI Resources, NIAID; NR-54970) were seeded at 5e5 cells per well in 50 μL aliquots in half area Greiner 96-well plates (Greiner Bio-One; Cat. No. 675090) 24 h prior to performing the neutralization assay. On separate U-bottom plates, patient plasma was plated in duplicates and serially 5-fold diluted in infection media (2% FBS, 1% glutamine, 1% sodium pyruvate in 1X DMEM) for a final volume of 28 μL per well. We also included ‘virus only’ and ‘media only’ controls. Twenty-five microliters containing ∼500 fluorescent forming units (FFUs) of a VSV encoding eGFP gene pseudotyped with one spike variant and ∼500 FFUs of a second VSV encoding an mCherry red gene pseudotyped with another spike variant was added to each well and incubated at 37°C. Prior to infection, Vero E6-TMPRSS2-T2A-ACE2 cells were washed with 1X PBS and then 50 μL of the incubated pseudotyped particles and patient plasma mixture was then transferred from the U-bottom 96-well dilution plates onto the monolayer and placed into an incubator at 37°C and 5% CO2. At 17 to 24 h post-incubation, the number of GFP- and RFP-expressing cells indicating viral infection were quantified using a Celigo Image Cytometer (Nexcelcom Bioscience). We first calculated the percent infection based on our ‘virus only’ controls and then calculate percent inhibition by subtracting the percent infection from 100. A non-linear curve and the half-maximal pseudoparticle neutralization titer (pNT50) were generated using GraphPad Prism.

## Data Availability

All data produced in the present study are available upon reasonable request to the authors

## Acknowledgments

We thank Stanford CTRU Biobank, Catherine A. Blish, Hector Bonilla, Karen Jacobson, Diego Martinez Mori, Kattria van der Ploeg, Sharon Chinthrajah, Monali Manohar, Tina Sindher, Will Collins, James Liu, Joe G, Anthony Buccanco, Katia Tkachenko, Mihir Shah, Allie Lee, Kathleen Jia, Eric Smith, Iris Chang, Evan Do and Diane Dunham for support with clinical protocol, patient care, and/or collection/provision of patient samples. We thank Lin Li, Christine Lin, Unurzul Jigmeddagva, Lakshmi Warrier, Veronica Gonzalez, and Emilia Basilio for assistance with participant recruitment, and sample collection/processing. We thank Christopher Wirblich and Matthias J. Schnell at Thomas Jefferson University for the VSV-ΔG-GFP/mCherry seed viruses.

## Supplemental information

Correspondence and requests for materials should be addressed to T.T.W and G.S.T

